# Long-term exposure to air-pollution and COVID-19 mortality in England: a hierarchical spatial analysis

**DOI:** 10.1101/2020.08.10.20171421

**Authors:** Garyfallos Konstantinoudis, Tullia Padellini, James Bennett, Bethan Davies, Majid Ezzati, Marta Blangiardo

## Abstract

**Background:** Recent studies suggested a link between long-term exposure to air-pollution and COVID-19 mortality. However, due to their ecological design, based on large spatial units, they neglect the strong localised air-pollution patterns, and potentially lead to inadequate confounding adjustment. We investigated the effect of long-term exposure to NO_2_ and PM_2·5_ on COVID-19 deaths up to June 30, 2020 in England using high geographical resolution. Methods We included 38 573 COVID-19 deaths up to June 30, 2020 at the Lower Layer Super Output Area level in England (n=32 844 small areas). We retrieved averaged NO_2_ and PM_2·5_ concentration during 2014-2018 from the Pollution Climate Mapping. We used Bayesian hierarchical models to quantify the effect of air-pollution while adjusting for a series of confounding and spatial autocorrelation.

**Findings:** We find a 0·5% (95% credible interval: −0·2%-1·2%) and 1·4% (−2·1%-5·1%) increase in COVID-19 mortality rate for every 1μg/m^3^ increase in NO_2_ and PM_2·5_ respectively, after adjusting for confounding and spatial autocorrelation. This corresponds to a posterior probability of a positive effect of 0·93 and 0·78 respectively. The spatial relative risk at LSOA level revealed strong patterns, similar for the different pollutants. This potentially captures the spread of the disease during the first wave of the epidemic.

**Interpretation:** Our study provides some evidence of an effect of long-term NO_2_ exposure on COVID-19 mortality, while the effect of PM_2·5_ remains more uncertain.

**Funding:** Medical Research Council, Wellcome Trust, Environmental Protection Agency and National Institutes of Health.

## Introduction

As of 30^th^ of June 2020, COVID-19 has caused more than 500 000 deaths globally, with an estimated case fatality of 1-4%.^1^ The UK is one of the countries most affected, with the an estimated 49 200 (44 700-53 300) more deaths than it would be expected from mid-February to 8^th^ May 2020 had the pandemic not taken place.^2^ Established risk factors of COVID-19 mortality include age, sex and ethnicity.^3^ Previous studies have observed a correlation between pre-existing conditions such as stroke, hypertension and diabetes.^4,5^ Long-term exposure to air-pollution has been hypothesised to worsen COVID-19 prognosis: either directly, as it can suppress early immune responses to the infection,^6^ or indirectly, as it can increase the risk of stroke, hypertension and other pre-existing conditions.^7,8^ Little is known about the effect of long-term exposure to air-pollution on COVID-19 mortality and evidence so far relies on ecological studies based on large areas. A study in the US, at county level, reported an 8% (95% confidence intervals: 2%-15%) increase in the COVID-19 death rate, for an increase of 1 μg/m^3^ in the long-term exposure to PM_2·5_ (atmospheric particulate matter that has a diameter of less than 25 micrometers).^6^ Another study in the US, at county level, examined the long-term effect of NO_2_, PM_2·5_ and O_3_ on COVID-19 case fatality (proportion of deaths among infected) and mortality rate and reported a 7·1% (1·2%-13·4%) and 11·2% (3·4%-19·5%) increase per 4·5ppb increase in NO_2_ for case fatality and mortality rate respectively.^9^ The same study reported weak evidence of an association between COVID-19 case fatality or mortality with long term exposure to PM_2·5_ and O3. A study in the Netherlands using municipalities reported that every unit increase in the long-term exposure to PM_2·5_, NO_2_ and SO_2_ was associated with 0·35, 2·3 and 1·8 additional COVID-19 deaths respectively.^10^ A study in England reported a significant association between long-term exposure to NO_2_, NO and O_3_ and COVID-19 deaths at Lower Tier Local Authorities (LTLA).^11^

Several methodological shortcomings limit the interpretability of previous studies:

1. They were based on data aggregated on large spatial units and thus suffer from ecological fallacy (grouped levels association do not reflect individual ones).^12^
2. Air pollution is characterised by high spatial variability, making the availability of mortality data at the same high spatial resolution crucial. In addition, a coarse geographical resolution might lead to inadequate adjustment for confounders, when these are available at higher resolution.
3. Most previous studies assessed cumulative deaths until mid or end of April and thus the generalisability of the results is limited to the early stages of the epidemic.^6,9,11^ Only one study had data available up to 5^th^ June 2020 capturing almost the entire first wave.^10^

In this nationwide study in England, we investigated the effect of long-term exposure to air pollution on COVID-19 mortality during the entire first wave of the epidemic, after accounting for confounding and spatial autocorrelation. We focused on exposure to NO_2_ and PM_2·5_. We downscaled the LTLA geographical information to the Lower Layer Super Output Area (LSOA) to alleviate the effect of ecological bias and exploit the variability of the exposure at high geographical resolution. We hypothesise that long-term exposure to these compounds worsens the prognosis of COVID-19 patients, as exposure to pollution can suppress early immune responses to the infection, leading to later increases in inflammation^6^ and as it can affect the onset of pre-existing conditions.^13–16^

## Methods

### Study population

We included all COVID-19 deaths up to June 30, 2020 in England as retrieved from Public Health England (PHE). For each death, PHE records individual data on age, sex and ethnicity, as well as the LTLA of the residential address. Information for the general population about age and sex is available from the Office of National Statistics (ONS) for 2018, whereas ethnicity is obtained from the 2011 census at the LSOA level.

We downscaled the LTLA geographical information to the LSOA level using census based weights to match the age, sex and ethnic composition of the deaths in each LTLA with that of the corresponding LSOAs. For more information about the downscaling procedure see Supplementary Material S1.1.

### Exposure

We considered exposure to NO_2_ and PM_2·5_ as indicators of air pollution. We selected these pollutants because: 1) they reflect different sources of air-pollution (NO_2_ reflects traffic related air-pollution, whereas PM_2·5_ is a combination of traffic and non-traffic sources), 2) they were considered in previous studies^6,9–11^ and 3) they are responsible for the highest number of years of life lost compared to other pollutants in Europe.^17^ We retrieved NO_2_ and PM_2·5_ concentration in England from the Pollution Climate Mapping (PCM; https://uk-air.defra.gov.uk/). The PCM produces annual estimates during 2001-2018 for NO_2_ and 2002-2018 for PM_2·5_ at 1×1km resolution for the UK. The PCM model is calibrated using monitoring stations across the nation and has high predictive accuracy, R^2^ =0·88 for NO_2_ and R^2^ = 0·63 for PM_2·5_.^18^ We defined long-term exposure to these compounds as the mean of the past 5 years for which data was available, i.e. 2014-2018. We weighted the exposure using a combination of population estimates available from the fourth version of Gridded Population of the World (GPW) collection at 1×1km grid as of 2020^19^ and from ONS at LSOA level as of 2018. For more information about the population weights see Supplementary Material S1.2.

### Confounders

We considered confounders related with meteorology, socio-demographics, disease spread, healthcare provision and health related variables (Table 1). As meteorological confounders, we considered temperature and relative humidity and calculated the mean for March-June 2018 as this is the latest year with data available at 1×1km grid retrieved from the MetOffice. We weighted temperature and relative humidity using the population weights calculated for the air-pollution exposure. As socio-demographical confounders we considered age, sex, 5ethnicity, deprivation, urbanicity, population density and occupation. Information on age (2018), sex (2018), ethnicity (2011), urbanicity (2011) and population density (2018) was available at the LSOA level from ONS. To adjust for deprivation, we used quintiles of the index of multiple deprivation at LSOA level in 2011 (Ministry of Housing, Communities and Local Government), excluding the dimension related to air quality. We used estimates of occupational exposures to COVID-19, as calculated by ONS, to adjust for high risk exposure to COVID-19, defined as those with a score higher than 80/100 (corresponding to at least >1 per week exposed to someone infected, Supplementary Material S1.3 and Table S1). To account for disease progression, we used the number of days since the 1st reported case and the number of positive cases in each LTLA (as of 30^th^ of June, as retrieved from PHE). For healthcare provision, we used the number of intensive care unit beds per population, in February 2020 per NHS trust, as retrieved by NHS. Last, as health-related variables, we considered smoking and obesity prevalence at the GP practice level during 2018-2019, as retrieved by PHE (Supplementary Material S1.3).

**Table 1.** Data sources used in the analysis

| Confounders | Source | Spatial Resolution | Temporal Resolution | Type |
| --- | --- | --- | --- | --- |
| Temperature | MetOffice<br>https://www.metoffice.gov.uk/ | 1km<sup>2</sup> | Mar-June 2018 | continuous |
| Relative humidity | MetOffice<br>https://www.metoffice.gov.uk/ | 1km<sup>2</sup> | Mar-June 2018 | continuous |
| Index of Multiple Deprivation | Ministry of Housing, Communities and Local Government<br>https://www.gov.uk/ | Lower layer super output area | 2011 | rank (quintiles) |
| Urbanicity | Office of National Statistics<br>https://www.ons.gov.uk/ | Lower layer super output area | 2011 | urban/rural |
| Days since 1<sup>st</sup> reported case | Public Health England | Lower tier local authority | Until 30<sup>th</sup> June | continuous |
| Number of positive cases | Public Health England | Lower tier local authority | Until 30<sup>th</sup> June | discrete (counts) |
| Population density | Office of National Statistics<br>https://www.ons.gov.uk/ | Lower layer super output area | 2018 | continuous (log transformed) |
| Number of intensive care unit beds | National Health Service<br>https://www.england.nhs.uk/ | National Health Service trust | February 2019 | continuous (per population) |
| Smoking | Public Health England<br>https://fingertips.phe.org.uk/ | General practitioner catchment area | 2018-2019 | continuous (prevalence) |
| Obesity | Public Health England<br>https://fingertips.phe.org.uk/ | General practitioner catchment area | 2018-2019 | continuous (prevalence) |
| High Risk Occupation | Office of National Statistics<br>https://www.ons.gov.uk/ | Middle layer super output area | 2011 | continuous (prevalence) |

### Statistical methods

We specified Bayesian hierarchical Poisson log-linear models to investigate the association of COVID-19 deaths and NO_2_ and PM_2·5_ independently. Spatial autocorrelation was modelled using a re-parametrisation of the Besag-York-Molliè conditional autoregressive prior distribution.^20,21^ We fitted four models including: 1) each pollutant (model 1), 2) each pollutant and the spatial autocorrelation term (model 2), 3) each pollutant and all confounders (model 3) and 4) each pollutant, the spatial autocorrelation term and all confounders (model 4). All models were adjusted for age, sex and ethnicity using indirect standardisation. In order to propagate the uncertainty resulted from the sampling we used for the downscaling (Supplementary Material S1.1), we fitted the models over 100 downscaled samples and then performed Bayesian model averaging to combine the estimates.^22^ We report results as posterior median of mortality relative risk for every 1 μg/m^3^ increase in the air-pollutants, 95% credibility intervals (CrI) and posterior probability that the estimated effect is positive. We also report posterior median of spatial mortality relative risks (exponential of the spatial autocorrelation term) and posterior probabilities that the spatial relative risks are larger than 1. The full model and prior specifications are given in the Supplementary Material S1.4. All models are fitted in INLA.^23^ Data and code are available on github (https://github.com/gkonstantinoudis/COVID19AirpollutionEn).

### Sensitivity analyses

We performed a series of sensitivity analyses. First, we repeated the main analyses using data at the LTLA level with all exposures and confounding weighted by population. Second, we examined if there is a differential effect of long-term exposure to air-pollution at the early stages of the epidemic, considering the lockdown (23^rd^ of March 2020) as a landmark. Third, we assessed the correlation between the latent field of the full model (model 4) with that of the model excluding or including only covariates indicating disease spread (i.e. number of tested positive cases and days since first reported cases). Fourth, we categorised pollutants into quintiles to allow more flexible fits. Fifth, we repeated the analysis using suspected cases as the outcome.

## Results

### Study Population

We identified 38 573 COVID-19 deaths with a laboratory confirmed test in England between 2^nd^ March and 30^th^ June (Figure 1). The age, sex and ethnicity distribution of the deaths follows patterns reported previously (Supplementary Material Tables S2-3).

**Fig 1.**
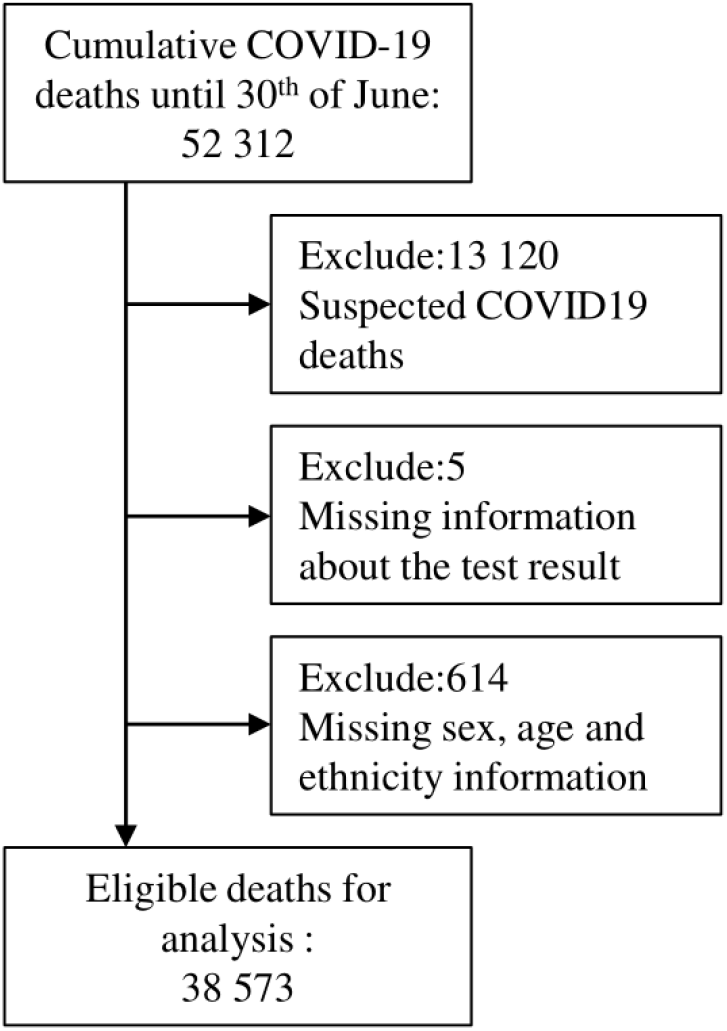
Flowchart of the COVID-19 deaths.

### Exposure

Figure 2 shows the population weighted air-pollutants at LSOA level in England. We observe that the localised variation of NO_2_, for instance due to the highways, is adequately captured at the spatial resolution of the LSOAs. The mean of NO_2_ is 16·17μg/m^3^ and it varies from 2·99μg/m^3^ in highly rural areas to 50·69μg/m^3^ in the big urban centres (Figure 2). The mean of PM_2·5_ is 9·84μg/m^3^ with a smaller variation, 5·14-14·22 μg/m^3^ (Figure 2).

**Fig 2.**
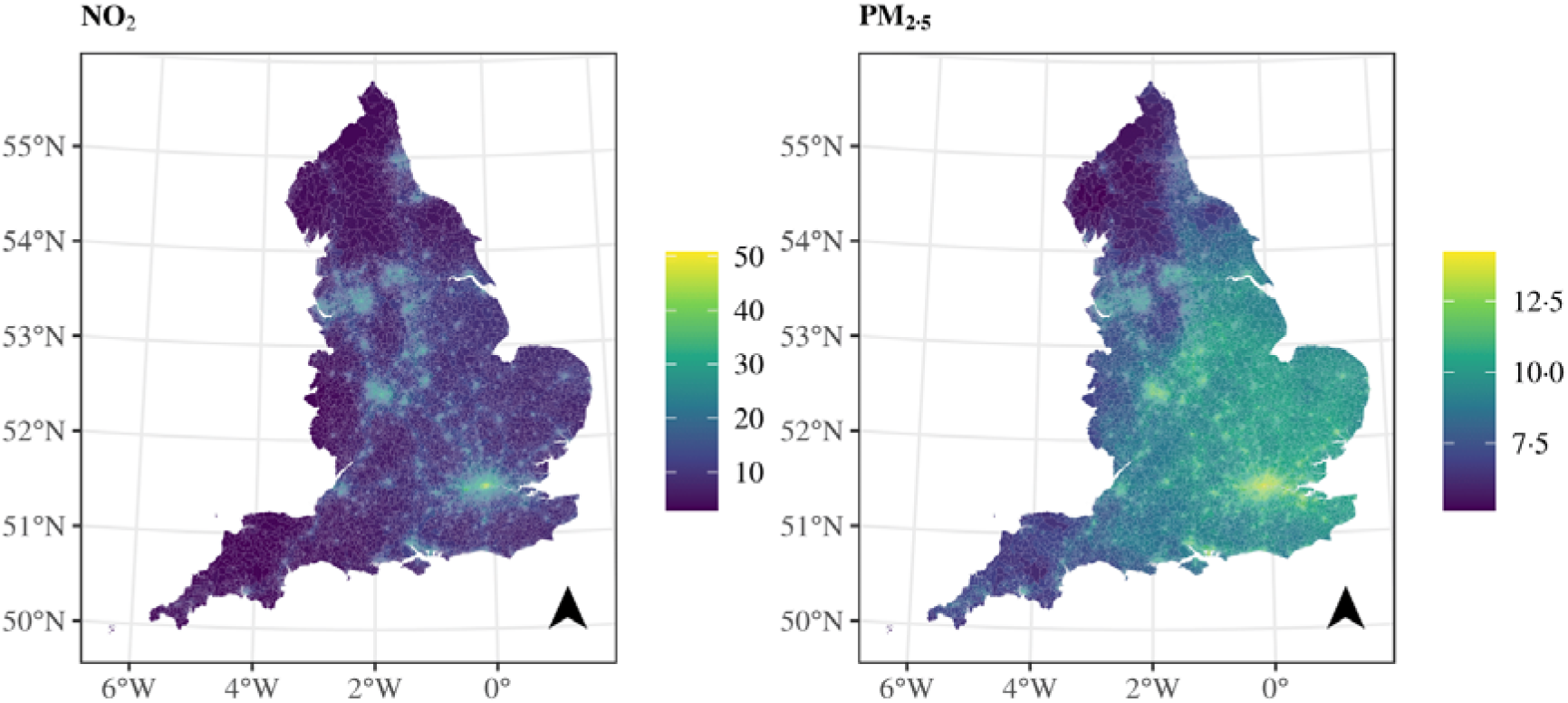
Population weighted exposure per LSOA.

### Confounders

Plots and maps of the confounders can be found in Supplementary Material, Figures S1-11.

### NO_2_

We observe a 2·6% (95%CrI: 2·4%-2·7%) increase in the COVID-19 mortality rate for every 1μg/m^3^ increase in the long-term exposure to NO_2_, based on model 1 (Figure 3 & Supplementary Material Table S4). There is still evidence of an effect, albeit smaller, once we adjust for spatial autocorrelation or confounders, with increases in the long-term exposure to NO_2_ of, respectively, 1·3% (0·8% - 1·8%), 1·8% (1·5% - 2·1%) for every 1 μg/m^3^. When we adjust for both autocorrelation and confounders the evidence is less strong, with estimates of 0·5% (−0·2% - 1·2%) for every 1μg/m^3^ (Figure 3 & Supplementary Material Table S4) and posterior probability of a positive effect reaching 0·93. The spatial relative risk in England varies from 0·24 (0·08-0·69) to 2·09 (1·30-3·11) in model 2 and from 0·30 (0·10-0·84) to 187 (1·18-2·93) in model 4, implying that the confounders explain very little of the observed variation (Figure 3). The variation is more pronounced in the cities and suburban areas (with posterior probability higher than 1; Figure 3).

**Fig 3.**
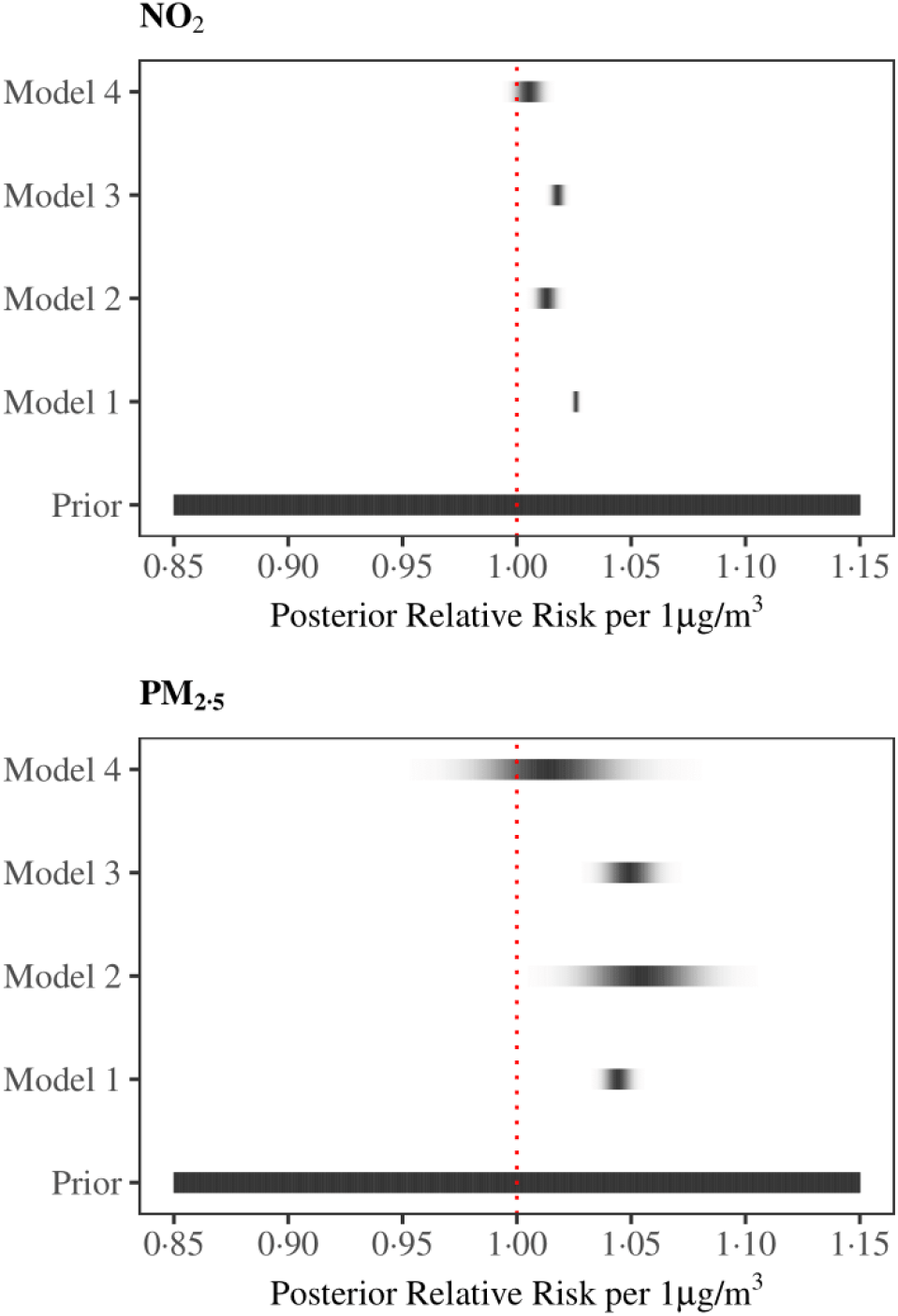
Density strips for the posterior of COVID-19 mortality relative risk with 1μg/m^3^ increase in NO_2_ (top panel) and PM_2·5_ (botom panel) averaged long-term exposure.

### PM_2·5_

We observe a 4·4% (3·7%-5·1%) increase in the mortality rate for every 1μg/m^3^ increase in the long-term exposure to PM_2·5_, based on model 1 (Figure 3 & Supplementary Material Table S5). When we adjust for spatial autocorrelation the effect increases slightly but the credibility intervals are wider, 5 4% (2·5%-8·4%), whereas it is similar when we adjust for confounding 4·9% (3·7%-6·2%) (Figure 3 & Supplementary Material Table S5). The effect is weak when we account for confounders and spatial autocorrelation 1·4% (−2·1%-5·1%) (Figure 3 & Supplementary Material Table S5). The posterior probability of a positive effect is lower than observed for NO_2_, and equal to 0·78. The spatial relative risk follows similar patterns as the one reported in the models for NO_2_, with the posterior median relative risk varying from 0·24 (0·12-0·46) to 2·26 (1·32-3·85) in model 2 and from 0·30 (0·15-0·57) to 190 (1·14-3·17) in model 4 (Supplementary Material Figure S12).

### Sensitivity Analyses

When LTLAs are the main geographical unit for analysis, the results are consistent, but higher in magnitude, potentially due to inadequate covariate and spatial autocorrelation adjustment due to the coarse geographical resolution (Supplementary Material Tables S6-7, Figures S13-14). Restricting the study period to March 23, 2020 (N=698) also results in similar estimates for both pollutants, however the uncertainty is higher (Supplementary Material Tables S8-9, Figures S15-16). The latent field of model 4, with NO_2_ as the pollutant, is similar to the latent fields of the models with and without the disease progression variables, with a correlation coefficient of 0·94 and 0·93 respectively (Supplementary Material Figure S17). The use of quintiles of the pollutants justifies the linearity assumption (Supplementary Material Figure S18). Finally, the results are consistent, but the evidence weaker, when suspected COVID-19 deaths are used instead (Supplementary Material Tables S10-11, Figures S19-20).

### Post-hoc analysis

In a post-hoc analysis we investigated if the evidence of an effect of NO_2_ on COVID-19 mortality can be attributed to pre-existing conditions. We selected hypertension, chronic obstructive pulmonary disease (COPD) and diabetes, because of 1) indications of previous literature that they increase the COVID-19 mortality risk,^4,5^ 2) previous literature that suggest an effect with long-term exposure NO_2_^14–16^ and 3) data availability. We retrieved prevalence data for these pre-existing conditions from PHE available at the GP practice level during 2018-2019 (https://fingertips.phe.org.uk/), Supplementary Material Figures S21-23. The effect of NO_2_ remain similar, 06% (−0·1% - 1·3%) with the posterior probability being 0·94 whereas the spatial relative risk highlights the same geographical locations, Supplementary Material Figure S24.

## Discussion

### Main findings

This is the first nationwide study in England investigating the effect of long-term exposure to NO_2_ and PM_2·5_ during 2014-2018 on COVID-19 mortality at LSOA level. The unadjusted models indicate that for every 1μg/m^3^ increase in the long-term exposure to NO_2_ and PM_2·5_ the COVID-19 mortality rates increase. After considering the effect of confounding and spatial autocorrelation the evidence is less strong for NO_2_, while for PM_2_._5_ there is larger uncertainty. The spatial relative risk has strong spatial patterns, identical for the different pollutants, potentially highlighting the effect of disease spread.

### Comparison with previous studies focusing on NO_2_

Our study is comparable with three previous studies in the US, England and the Netherlands assessing the long-term effect of NO_2_ in COVID-19 mortality. The study in the US focused on deaths reported by April 29, 2020, using 3·122 counties. For the exposure, they calculated the mean of daily concentrations during 2010-2016 as modelled by a previously described ensemble machine learning model (R^2^=0·79).^25^ They reported a 7·1% (1·2%-13·4%) increase in mortality per 4·5ppb (1ppb=1·25μg/m^3^) increase in NO_2_ after adjusting for confounders and spatial autocorrelation^9^ (that is approximately 1·3% increase per 1 (μg/m^3^). The study in England, with partly overlapping data as in our analysis, also reported a significant association between NO_2_ and COVID-19 mortality (p<0·05). For the analysis they focused on COVID-19 deaths reported in England up to April 10, 2020, used 317 LTLAs, and did not account for spatial autocorrelation.^11^ The study in the Netherlands using 335 municipalities and mean exposure during 2015-2019 reported 0·35 (0·04-0·66) additional COVID-19 deaths for every 1μg/m^3^ increase in NO_2_ after adjusting for confounders and certain spatial controls, such as transmission beyond the Dutch national borders^10^. Since the mean number of deaths in their sample is 16·86, the above estimate translates to a 2·0% increase in the COVID-19 mortality for every 1μg/m^3^ increase in NO_2_.

### Comparison with previous studies focusing on PM_2·5_

Our study is comparable with previous studies assessing the long-term effect of PM_2·5_ on COVID-19 mortality. The aforementioned study in the US also assessed the effect of PM_2·5_ on COVID-19 mortality.^9^ Their exposure model was previously validated having an R^2^ = 0·89 for the annual estimates.^26^ The evidence for PM_2·5_ was weak, namely 10·8% (−1·1-24·1%) per 3·4μg/m^3^ increase in PM_2·5_ concentration (that is approximately 3·2% increase per 1μg/m^3^) after adjusting for confounding and spatial autocorrelation. Our study comes in contrast with another study in the US that used deaths reported until April 22^nd^, 2020 and counties as the geographical unit.^6^ For the exposure, they used previously validated monthly PM_2·5_concentrations (R^2^= 0·70)^27^ and averaged them during 2000 and 2016. After adjusting for confounding but not for spatial autocorrelation, they found an 8% (2%-15%) increase in the COVID-19 death rate for an increase of 1μg/m^3^ in PM_2·5_ concentration. Our study comes also in contrast with the study in the Netherlands that reported 2·3 (1·3-3·0) additional COVID-19 deaths for an increase of 1μg/m^3^ in the averaged long-term PM_2·5_ concentration.^10^ Having a mean number of deaths equal to 16·86, the above estimate translates to a 13·6% increase in the COVID-19 mortality rate for an increase of 1μg/m^3^ in PM_2·5_ concentration.

### Strengths and Limitations

Our study is the first study that examines the association between long-term exposure to NO_2_ and PM_2·5_ at very high geographical precision. The spatial unit of our analysis is LSOAs, for which there are 32 844 in England (~130 000km), whereas previous studies have used 317 LTLAs in England, counties in the US (3 122 in an area ~9·8 million km^2^) and municipalities in the Netherlands (334 in an area ~41 500km^2^). Such high-resolution allows capturing the localised geographical patterns of the pollutants but also ensures adequate confounding and spatial autocorrelation adjustment. Our study also covers, so far, the largest temporal window of the epidemic (capturing the entire first wave, Figure S25 Supplementary Material), while most previous studies focused on the early to mid-stages of the first wave. This ensures better generalisability of the results. We also adjusted for spatial autocorrelation, which was found to be a crucial component in the model. Not accounting for spatial autocorrelation, when spatial autocorrelation is present, is expected to give rise to narrower credible intervals and false positive effects.^28^

Our study has also some limitations. The downscaling procedure will likely inflate the reported credible intervals. However, this naturally reflects the uncertainty of the place of residence resulted from the downscaling approach. Although we consider small areas, the study is still an ecological one and thus the reported effects do not reflect individual associations.^12^ Case fatality might have been a more appropriate metric for the analysis, since disease spread is accounted for in the denominator. Nevertheless, given the asymptomatic infections and the fact that number of reported infections is not a random sample of the general population, the number of COVID-19 cases per LTLA is not reliable at this stage. However, part of the disease spread was captured in the spatial autocorrelation term. We did not account for population mobility during 2014-2018, and assumed constant residence and thus levels of exposure to air-pollution. We also could not account for non-residential air-pollution exposure.

### Interpretation

Compared to the previous studies, our results are the smallest in magnitude, likely because of the high geographical precision that allows more accurate confounding and spatial autocorrelation adjustment. In addition, we report the weakest evidence of an effect, which could be due to lack of power and individual exposure data. Nevertheless, as for NO_2_ we find a high posterior probability of an effect on mortality, we argue that a potential explanation might be the mediation effect of pre-existing conditions. While in our analysis the inclusion of area-level prevalence of hypertension, diabetes and COPD did not change the results, the ecological nature of the pre-existing conditions data does not allow us to account for the mediation effect at the individual level. None of the previous studies have accounted for pre-existing conditions. Similarly, the weak, but positive, effect observed for PM_2·5_ could be an attribute to pre-existing conditions, or even disease spread, as recent studies have suggested that PM_2·5_ can proliferate COVID-19 transmission.^29^

Our analysis captured strong spatial autocorrelation. The observed pattern could reflect residual variation from a potential inadequate covariate adjustment (including disease spread), spatial variation of pre-existing conditions, other unknown spatial confounders or a combination from all above. In a sensitivity analysis, we observed that the factors associated with disease transmission left the latent field unchanged (Supplementary Material Figure S17), as did the inclusion of hypertension, diabetes and COPD (Supplementary Material Figure S24). When we restricted the analysis to the pre-lockdown period, the latent field for both pollutants captured London and Birmingham, i.e. the cities with the first outbreaks. Considering the above, and the fact that COVID-19 is an infectious disease, we believe that large variation of Figure 4 is likely due to disease spread, which is not adequately captured in the disease progression covariates.

**Fig 4.**
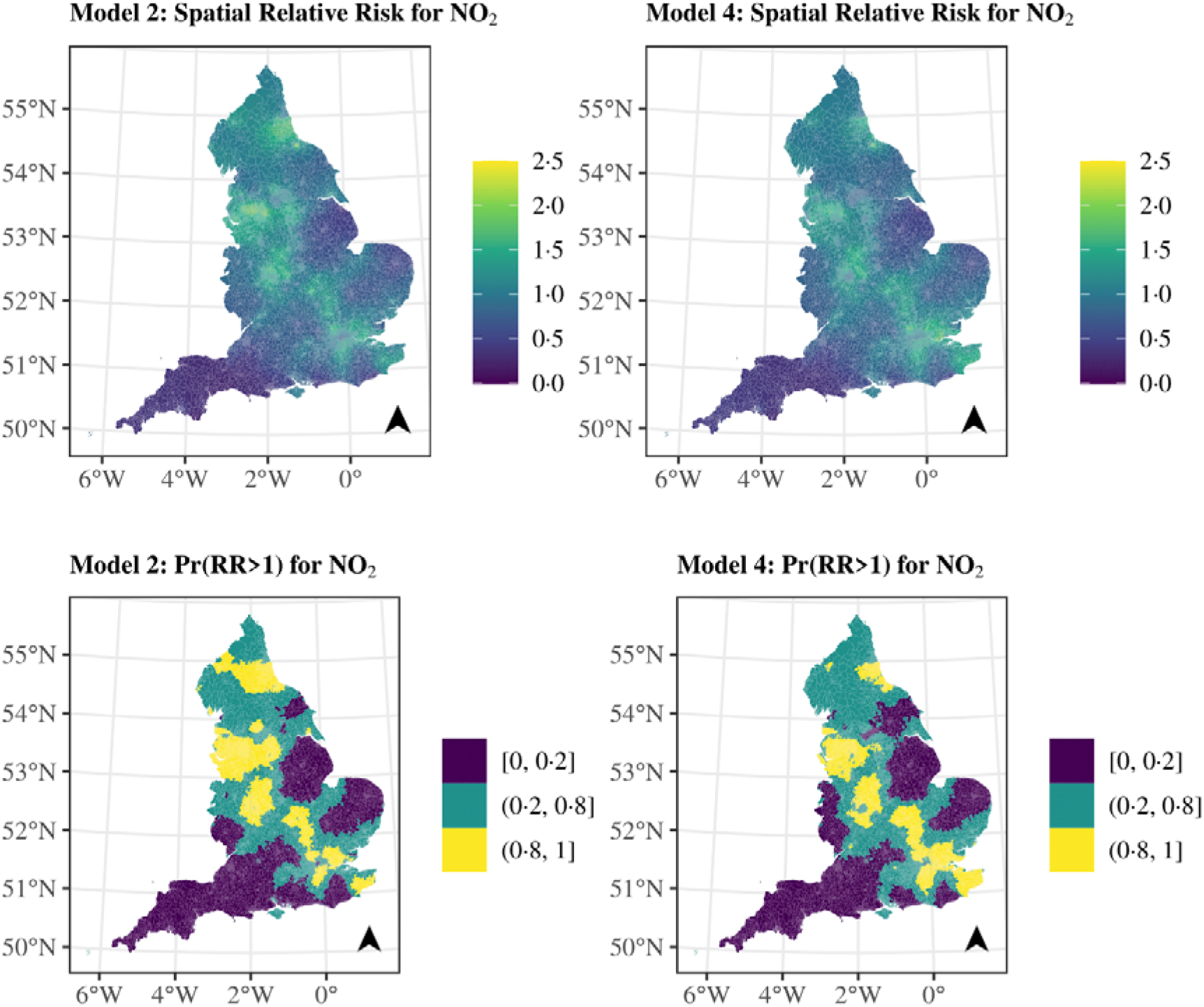
Median posterior spatial relative risk (exponential of the spatial autocorrelation term) and posterior probability that the spatial relative risk is larger than 1 for the models with NO_2_ and a spatial autocorrelation term and the fully adjusted NO_2_ model.

### Conclusion

Overall, this study provides some evidence of an association between averaged exposure during 2014-2018 to NO_2_ and COVID-19 mortality, while the role of PM_2·5_ remains more uncertain.

## Data Availability

Outcome data can be available after a request to PHE. Confounding data and the Rcode are online available on https://github.com/gkonstantinoudis/COVID19AirpollutionEn.

## Contributors

Conceptualisation: G.K., T.P., M.B.; Methodology: G.K., T.P., M.B.; Formal analysis: G.K., T.P.; Validation: G.K., T.P., J.B., B.D., M.E., and M.B.; Writing-original draft: G.K.; Writing-review and editing: G.K., T.P., J.B., B.D., M.E., and M.B.; Resources: G.K., M.B., M.E.; Supervision: M.B. All authors read and approved the final manuscript.

## Ethical approval

Only aggregated count data were used in this analysis; no individual data were used.

## Acknowledgements

G.K.is supported by an MRC Skills Development Fellowship [MR/T025352/1]. M.B and T.P are supported by a National Institutes of Health, grant number [R01HD092580-01A1]. J.B. and M.E. are supported by the Pathways to Equitable Healthy Cities grant from the Wellcome Trust [209376/Z/17/Z] and by a grant from the US Environmental Protection Agency (EPA), as part of the Centre for Clean Air Climate Solution (assistance agreement no. R835873). This article has not been formally reviewed by the EPA. The views expressed in this document are solely those of the authors and do not necessarily reflect those of the EPA. The EPA does not endorse any products or commercial services mentioned in this publication.

## References

1. Hauser A, Counotte MJ, Margossian CC, et al. Estimation of SARS-CoV-2 mortality during the early stages of an epidemic: a modelling study in Hubei, China and northern Italy. medRxiv 2020.

2. Kontis V, Bennett JE, Parks RM, et al. Age-and sex-specific total mortality impacts of the early weeks of the Covid-19 pandemic in England and Wales: Application of a Bayesian model ensemble to mortality statistics. medRxiv 2020.

3. Wu Z, McGoogan JM. Characteristics of and Important Lessons From the Coronavirus Disease 2019 (COVID-19) Outbreak in China: Summary of a Report of 72 - 314 Cases From the Chinese Center for Disease Control and Prevention. JAMA 2020; 323(13): 1239–42.

4. Williamson E, Walker AJ, Bhaskaran KJ, et al. OpenSAFELY: factors associated with COVID-19-related hospital death in the linked electronic health records of 17 million adult NHS patients. medRxiv 2020.

5. Yang J, Zheng Y, Gou X, et al. Prevalence of comorbidities and its effects in patients infected with SARS-CoV-2: a systematic review and meta-analysis. International Journal of Infectious Diseases 2020; 94: 91-5.

6. Wu X, Nethery RC, Sabath BM, Braun D, Dominici F. Exposure to air pollution and COVID-19 mortality in the United States. medRxiv 2020.

7. Giorgini P, Di Giosia P, Grassi D, Rubenfire M, D Brook R, Ferri C. Air pollution exposure and blood pressure: an updated review of the literature. Current pharmaceutical design 2016; 22(1): 28–51.

8. Scheers H, Jacobs L, Casas L, Nemery B, Nawrot TS. Long-term exposure to particulate matter air pollution is a risk factor for stroke: meta-analytical evidence. Stroke 2015; 46(11): 3058–66.

9. Liang D, Shi L, Zhao J, et al. Urban Air Pollution May Enhance COVID-19 Case-Fatality and Mortality Rates in the United States. medRxiv 2020.

10. Cole M, Ozgen C, Strobl E. Air Pollution Exposure and COVID-19. 2020.

11. Travaglio M, Yu Y, Popovic R, Leal NS, Martins LM. Links between air pollution and COVID-19 in England. *medRxiv* 2020.

12. Wakefield J. Ecologic studies revisited. Annu Rev Public Health 2008; 29: 75-90.

13. Andersen ZJ, Bønnelykke K, Hvidberg M, et al. Long-term exposure to air pollution and asthma hospitalisations in older adults: a cohort study. Thorax 2012; 67(1): 6–11.

14. Balti EV, Echouffo-Tcheugui JB, Yako YY, Kengne AP. Air pollution and risk of type 2 diabetes mellitus: a systematic review and meta-analysis. Diabetes research and clinical practice 2014; 106(2): 161–72.

15. Cai Y, Zhang B, Ke W, et al. Associations of short-term and long-term exposure to ambient air pollutants with hypertension: a systematic review and meta-analysis. Hypertension 2016; 68(1): 62–70.

16. Zhang Z, Wang J, Lu W. Exposure to nitrogen dioxide and chronic obstructive pulmonary disease (COPD) in adults: a systematic review and meta-analysis. Environmental Science and Pollution Research 2018; 25(15): 15133–45.

17. Ortiz A. Air Quality in Europe-2019 Report. European Environment Agency, Luxembourg 2019.

18. Brookes DS, John; Kent, Andrew; Whiting, Sally; Rose, Rebecca; Williams, Chris. Technical Report on UK Supplementary Assessment Under the Air Quality Directive (2008/50/EC), the Air Quality Framework Directive (96/62/EC) and Fourth Daughter Directive (2004/107/EC) for 2015. 2017.

19. Center for International Earth Science Information Network - CIESIN - Columbia University. Gridded Population of the World, Version 4 (GPWv4): Population Count, Revision 11. Palisades, NY: NASA Socioeconomic Data and Applications Center (SEDAC); 2018.

20. Besag J, York J, Mollié A. Bayesian image restoration, with two applications in spatial statistics. Annals of the institute of statistical mathematics 1991; 43(1): 1–20.

21. Simpson D, Rue H, Riebler A, Martins TG, Sørbye SH. Penalising model component complexity: A principled, practical approach to constructing priors. Statistical science 2017; 32(1): 1–28.

22. Gómez-Rubio V, Bivand RS, Rue H. Bayesian model averaging with the integrated nested Laplace approximation. Econometrics 2020; 8(2): 23.

23. Rue H, Martino S, Chopin N. Approximate Bayesian inference for latent Gaussian models by using integrated nested Laplace approximations. Journal of the royal statistical society: Series b (statistical methodology) 2009; 71(2): 319–92.

24. Linton NM, Kobayashi T, Yang Y, et al. Incubation period and other epidemiological characteristics of 2019 novel coronavirus infections with right truncation: a statistical analysis of publicly available case data. Journal of clinical medicine 2020; 9(2): 538.

25. Di Q, Amini H, Shi L, et al. Assessing NO_2_ Concentration and Model Uncertainty with High Spatiotemporal Resolution across the Contiguous United States Using Ensemble Model Averaging. Environmental science & technology 2019; 54(3): 1372–84.

26. Di Q, Amini H, Shi L, et al. An ensemble-based model of PM2. 5 concentration across the contiguous United States with high spatiotemporal resolution. Environment international 2019; 130: 104909.

27. Van Donkelaar A, Martin RV, Li C, Burnett RT. Regional estimates of chemical composition of fine particulate matter using a combined geoscience-statistical method with information from satellites, models, and monitors. Environmental science & technology 2019; 53(5): 2595–611.

28. Lee D, Sarran C. Controlling for unmeasured confounding and spatial misalignment in long-term air pollution and health studies. Environmetrics 2015; 26(7): 477–87.

29. Bianconi V, Bronzo P, Banach M, Sahebkar A, Mannarino MR, Pirro M. Particulate matter pollution and the COVID-19 outbreak: results from Italian regions and provinces. Archives of Medical Science 2020; 16(1).

